# A Gynecology-Led Transgender Clinic: Retrospective Review of Referral Patterns and Quality Indicators

**DOI:** 10.1101/2020.05.27.20111831

**Authors:** Jennifer McCall, Jennifer She, Jessica Pudwell, Maria Kielly, Ashley Waddington

## Abstract

**Background:** The Transgender Clinic at the Kingston Health Sciences Centre (KTC) was created in 2017 to meet the needs of transgender patients.

**Methods:** A retrospective review of all KTC patient charts was completed. The primary outcomes were the referral pattern to, and services provided by KTC. The secondary outcomes were the quality indicators i) rates of cervical screening and ii) offers of fertility preservation.

**Results:** 108 patients were referred and 92 were seen; the median age was 20 years old. 77.2% of all patients sought, and 72.8% ultimately received, hormone therapy. Cervical cancer screening was documented as up to date for 69.8% of eligible people. 81.7% of eligible patients had documented fertility preservation offers.

**Conclusion:** Accessible local transgender care is a standard that can and should be met in Canada and internationally.

- 77.2% of patients sought hormone therapy, which is in the scope of providers from many specialties
- Accessible local transgender care is a standard than can and should be met in Canada and internationally
- Gynecologists, pediatricians, and primary care providers are well-suited to provide transgender care

## Introduction

Transgender people have unique health needs and encounter many barriers to meeting these needs. ^1^ Barriers include lack of access to care due to refusal by health care providers, anticipation of stigma in health care, discrimination, expense, improper pronoun use, long wait times, expectations that the patient will take the lead in their care, and provider lack of knowledge. ^2–5^ When transgender people are unable to access care, the consequences can be dire. A large American survey found that 27.6% of transgender people who were unable to access care turned to substance use to cope. ^2^ Rotondi found that 26.8% of transgender Ontarians had used nonprescribed hormones and a small number had attempted self-performed gender-affirming surgeries. ^6^

A sample of interviewed Ontario physicians disclosed that accessing resources and confusing health system organization were barriers to competently providing transgender care. ^7^ Lack of medical knowledge has also been identified as a barrier. ^7–9^ These obstacles make it difficult for physicians to provide transgender care.

Transgender health clinics are mainly led by family physicians, endocrinologists, pediatricians, psychiatrists and psychologists. ^10–12^ Most clinics are multidisciplinary. ^13,14^ Some are structured with one specialty as the primary or initial assessor (often Psychology or Psychiatry) with subsequent referral to other specialists. ^11,12,15^

Transgender men and gender non-conforming individuals assigned female at birth are less likely than cisgender women to be up to date on cervical cancer screening (Absolute Odds Ratio 0.63 (CI 0.47, 0.85) and 0.33 (CI 0.14, 0.79), respectively). ^16,17^ Rainbow Health Ontario guidelines for transgender health care recommend that transgender men and gender non-conforming individuals with a cervix follow the same screening recommendations as cisgender women. ^18,19^ Ontario’s 2011 goal for cervical cancer screening is 85%; from 2009-2011 65% of cis-women were screened. ^20^ In 2015, Kingston’s late-to-screening rate for cervical cancer was 22.8-33.7%, which was lower than the provincial average of 36%. ^21^

Rainbow Health Ontario recommends a discussion of fertility preservation at every initial assessment prior to cross-hormone therapy initiation. ^18,22^ When desired, early referral is also recommended due to lengthy wait times for these services. In an Australian questionnaire of 409 transgender and non-binary adults, 95% felt that all transgender and non-binary people should be offered fertility preservation. ^23^ Despite this, uptake of fertility preservation in this same population was only 7% (self-reported). ^23^ This is consistent with chart reviews, which have documented interest in fertility preservation among pediatric and adolescent patients to be 3-12%. ^15^,^24^

### Clinic description

The Transgender Clinic (KTC) at the Kingston Health Sciences Centre (KHSC) was established in July 2017 by Dr. Ashley Waddington (AW). AW recognized a dearth of transgender-specific health care services in the Kingston community and created KTC to fill that gap. KTC is a gynecologist-led clinic serving all genders and ages in Kingston and the surrounding Southeast Local Health Integration Network. It runs one half-day per month. Kingston, Ontario is a mid-sized Canadian city with a population of 117,660. ^25^ It is the only major Canadian city in a 200km radius. KHSC is an academic centre affiliated with Queen’s University with a catchment area of approximately 500 000 people.

### Definitions

Unless specified, in this publication the term “transgender” refers to people who identify differently from their assigned sex at birth; it encompasses but is not limited to transgender, transsexual, two-spirit, non-binary, and gender non-conforming identities. Hormone therapy broadly refers to puberty blockers, hormonal methods to induce amenorrhea (e.g., intramuscular medroxyprogesterone acetate), contraception, puberty blockers (e.g. spironolactone for transgender females) and cross-hormone therapy (e.g., testosterone for transgender males and estrogen for transgender females). Hysterectomy is uniquely identified and includes any method of hysterectomy with or without salpingectomy and oophorectomy. Bottom surgery includes orchiectomy, penectomy, and genital reconstruction (e.g., vaginoplasty, phalloplasty). Top surgery refers to mastectomy with or without chest reconstruction and breast implantation. Other surgery refers to surgeries other than top or bottom surgery, such as voice masculinization and facial recontouring.

People considered eligible for cervical cancer screening were those with a cervix who met Cancer Care Ontario guidelines. Regarding fertility preservation, patients were considered ineligible if they were already on cross-hormone therapy or had undergone gonadectomy prior to their initial visit. For the purpose of this study, patients were not considered eligible for fertility preservation offer if neither cross-hormone therapy nor gonadectomy was being initiated (e.g., a patient seeking menstrual suppression using reversible contraceptive methods). These criteria were used as we were most interested in investigating the number of people for whom the opportunity to preserve fertility was missed when initiating therapies at our clinic.

### Objectives

The primary outcomes are the referral pattern to, and services provided by KTC. The secondary outcomes are the quality indicators i) rates of cervical screening and ii) offers of fertility preservation. The mental health history of patients in this clinic has been described elsewhere. ^26^

## Methods

A retrospective review of all KTC patient charts was completed in May 2019. KTC was established in July 2017, though some earlier records are included as they had been referred to AW’s General Gynecology clinic prior to the official opening of KTC. Patients were excluded if they had been discharged from KTC without ever being seen, either due to inability to connect with the patient to make an appointment or because the patient was referred elsewhere prior to a KTC visit.

### Ethics

This study was approved for ethical compliance by the Queen’s University Health Sciences and Affiliated Teaching Hospitals Research Ethics Board. Patient consent was not required to access and use these data for research purposes. To maintain confidentiality, any planned subgroup analysis with less than 6 patients was excluded.

### Data collection

Two abstractors were responsible for data collection. JM, an Obstetrics and Gynecology resident, and JS, a senior medical student. JM trained JS by reviewing 10 charts together in-person and subsequently responding to queries. A standardized data collection form was used including a standardized data dictionary for data reduction. JM reviewed all but one chart, including those reviewed by JS, to ensure high reliability between reviewers. The single chart not reviewed by JM belonged to a personal acquaintance of JM and patient confidentiality was prioritized. Data collected included assigned sex at birth, current gender identity, age at time of referral, whether the patient had been seen in KTC, referral source, reason for referral both per provider on the referral and per patient at clinic visits, treatments received, previous treatment, referrals out to other specialists, wait times to first appointment and to therapy, wait times to see other specialists, the month and year of referral, length of follow-up, lost to follow-up, status of cervical cancer screening, and fertility preservation offer.

### Data analysis

Data are summarized as count (percent) and median [interquartile range].

### Patient and Public Involvement

The outcome measures included patient’s preferred gender identity and measurement of the reasons the patients sought referral to the transgender health clinic as elicited over several clinic visits. Patients were not involved in the design of the study. Results will be disseminated to patients via posting a public-friendly abstract in the clinic. A key outcome of this study is that the clinic team became aware that a patient satisfaction survey would be a key component of care moving forward.

## Results

### Patient characteristics

One hundred and eight patients were referred to KTC prior to May 7, 2019 (Figure 1). One patient met exclusion criteria. Ninety-two patients had been seen in KTC by May 7, 2019; the remaining 15 had initial appointments pending. Patient characteristics are summarized in Table 1. Median age at time of referral was 20 years old [IQR 16-28]. Patients 15 years old or younger constituted 17.8% of the clinic population. The majority of patients were assigned female at birth (69.2%). Current male gender identity predominated (59.8%).

**Figure 1.**
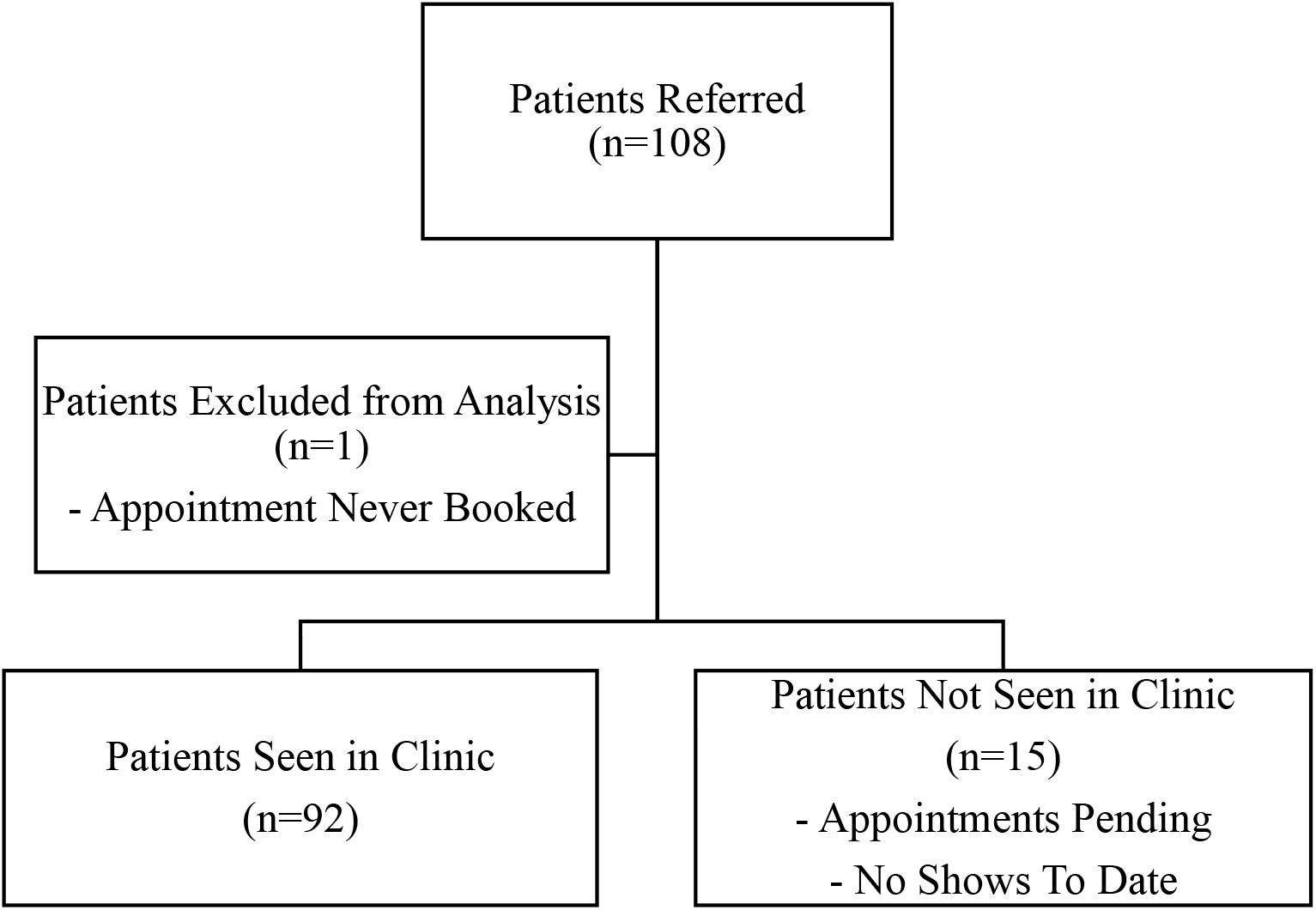
Study Population

**Table 1.**
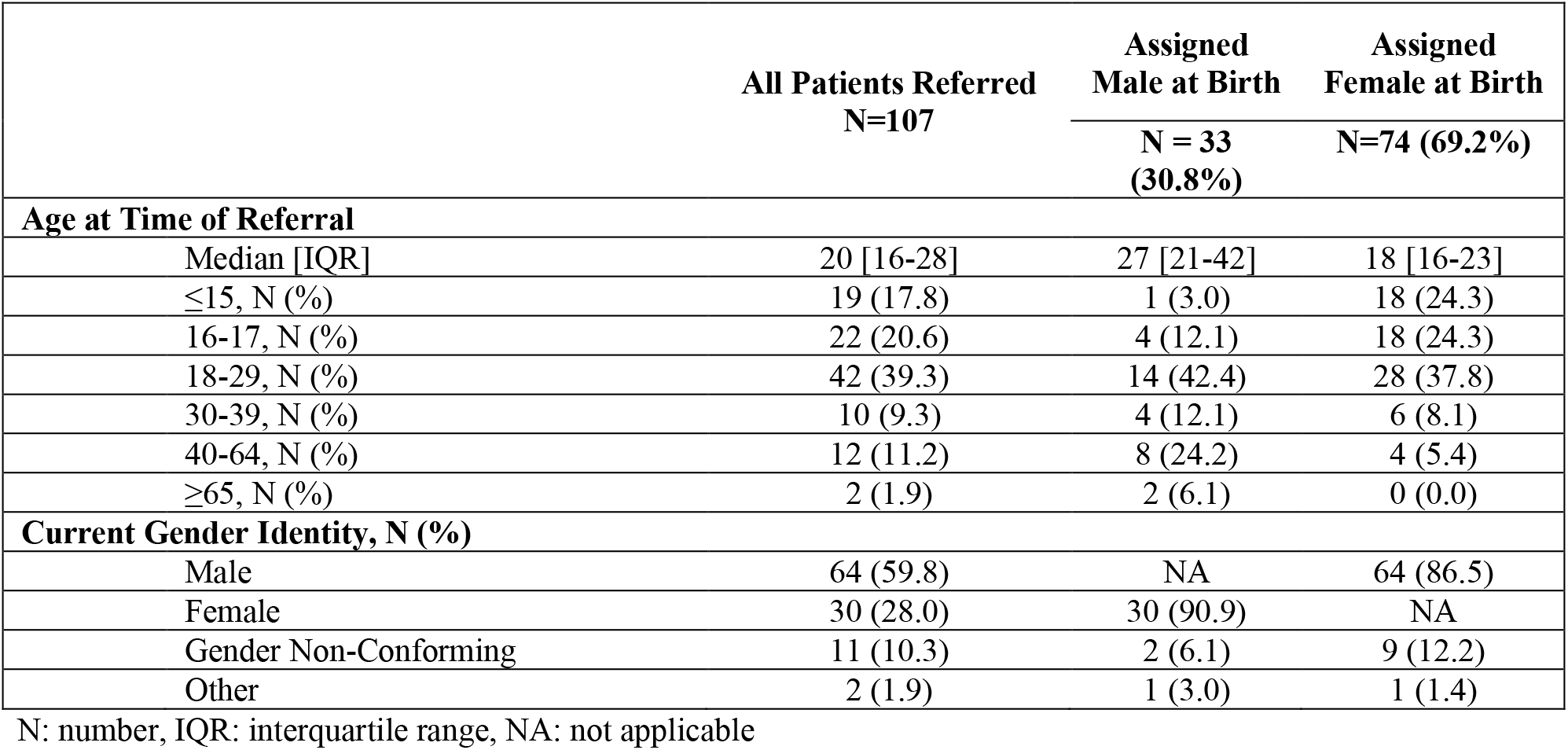
– Patient Characteristics – Age and Gender Identity.

|  | **All Patients Referred**<br>**N=107** | **Assigned**<br>**Male at Birth**<br>**N = 33**<br>**(30.8%)** | **Assigned**<br>**Female at Birth**<br>**N=74 (69.2%)** |
| --- | --- | --- | --- |
| **Age at Time of Referral** |  |  |  |
| Median [IQR] | 20 [16-28] | 27 [21-42] | 18 [16-23] |
| ≤15, N (%) | 19 (17.8) | 1 (3.0) | 18 (24.3) |
| 16-17, N (%) | 22 (20.6) | 4 (12.1) | 18 (24.3) |
| 18-29, N (%) | 42 (39.3) | 14 (42.4) | 28 (37.8) |
| 30-39, N (%) | 10 (9.3) | 4 (12.1) | 6 (8.1) |
| 40-64, N (%) | 12 (11.2) | 8 (24.2) | 4 (5.4) |
| ≥65, N (%) | 2 (1.9) | 2 (6.1) | 0 (0.0) |
| **Current Gender Identity, N (%)** |  |  |  |
| Male | 64 (59.8) | NA | 64 (86.5) |
| Female | 30 (28.0) | 30 (90.9) | NA |
| Gender Non-Conforming | 11 (10.3) | 2 (6.1) | 9 (12.2) |
| Other | 2 (1.9) | 1 (3.0) | 1 (1.4) |

N: number, IQR: interquartile range, NA: not applicable

### Referrals

Referral information is summarized in Table 2. Primary care providers made most of the referrals (64.5%) though specialist pediatricians also made a significant contribution (15.9%). The majority of referrals were for cross hormone therapy (48.6% according to referring provider and 77.2% according to the patient in clinic). A significant number of patients sought top surgery during their care at KTC (46.7%), though this was only identified in 5.6% of provider referrals. Referring providers included all the reasons a patient wished to be referred in only 12% of cases. However, all referring providers indicated at least one reason for referral that aligned with their patient’s intentions.

**Table 2.**
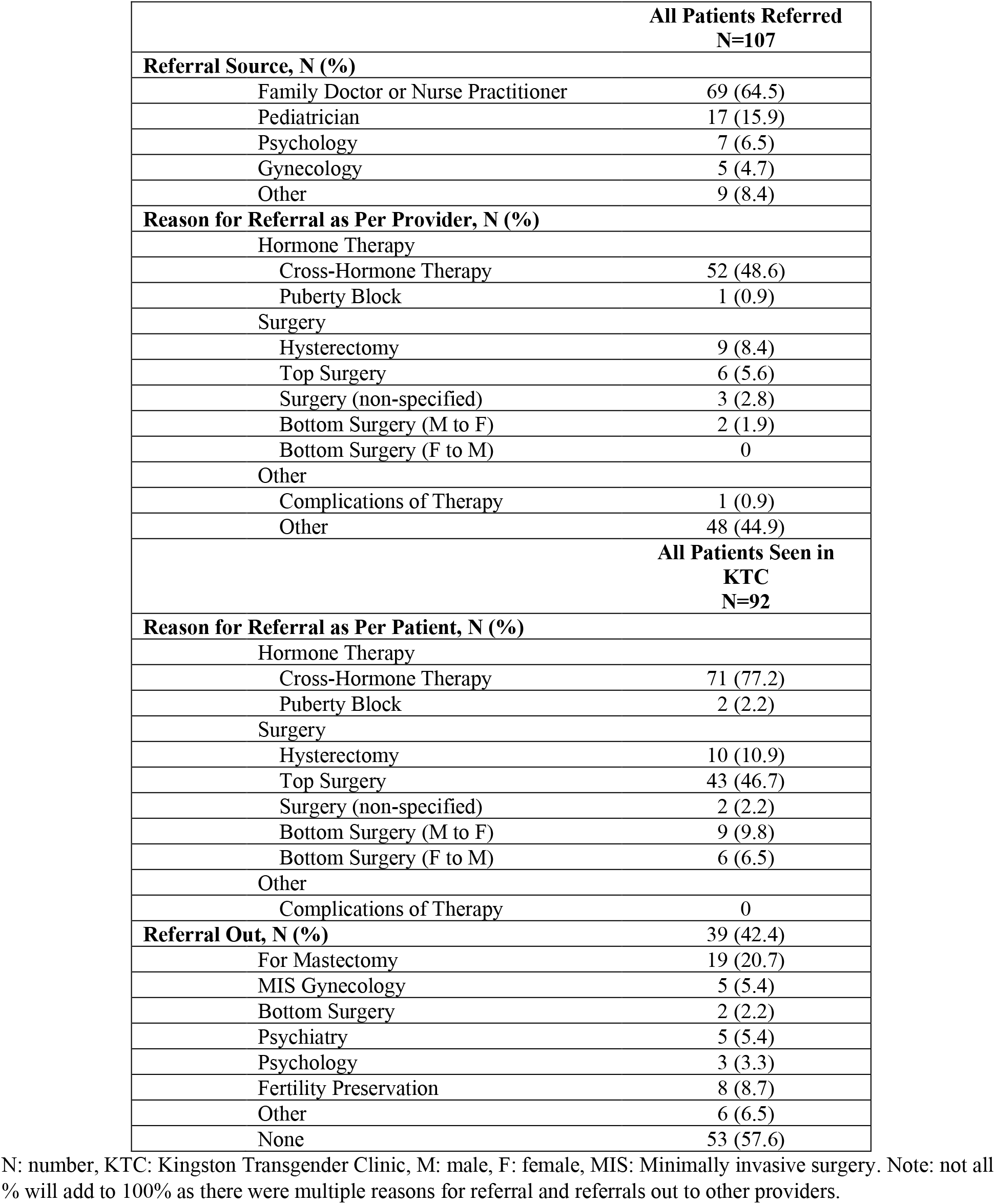
– Referrals Received and Requested – Requesting/Receiving Providers and Reasons for Referral to KTC.

Thirty-nine patients required referral beyond KTC (42.4%). Of these referrals, most were for mastectomy (48.7%). Reproductive Endocrinology and Infertility for fertility options, Psychiatry, and Minimally Invasive Surgery (MIS)-Gynecology were also consulted for a significant number of these referrals (20.5%, 12.8%, and 12.8%, respectively). Eight of forty-nine (16.3%) patients who were offered fertility preservation accepted a referral to Reproductive Endocrinology and Infertility.

### Treatments

At the time of their initial visit to KTC, 40.2% of patients had already received hormone therapy. Through KTC, 77.2% of all patients sought, and 72.8% ultimately received, hormone therapy. Menstrual suppression was received by 11.9% of patients and 4.3% underwent puberty blockade. Regarding surgery, 46.7% of patients sought top surgery and 21.7% were ultimately referred.

Ninety-five percent of top surgeries were mastectomies (data not shown). Only 4 of 63 assigned female at birth patients received hysterectomy (6.3%).

### Wait times

Wait times steadily increased from the date that KTC opened (Figure 2A). Patients referred in the three months prior to the first clinic (April-June 2017) waited a median of 58 days [IQR 47-85], while patients referred one year later (April-June 2018) waited a median of 264 days [IQR 231-291]. This mirrored the increase in the number of referrals (Figure 2B). In the three-month period prior to the first clinic, KTC received 10 referrals. Over the first year of operations, KTC received 16-19 referrals per 3-month period. The median wait time of all patients who had an initial visit to KTC was 207 days [IQR 176-238].

**Figure 2.**
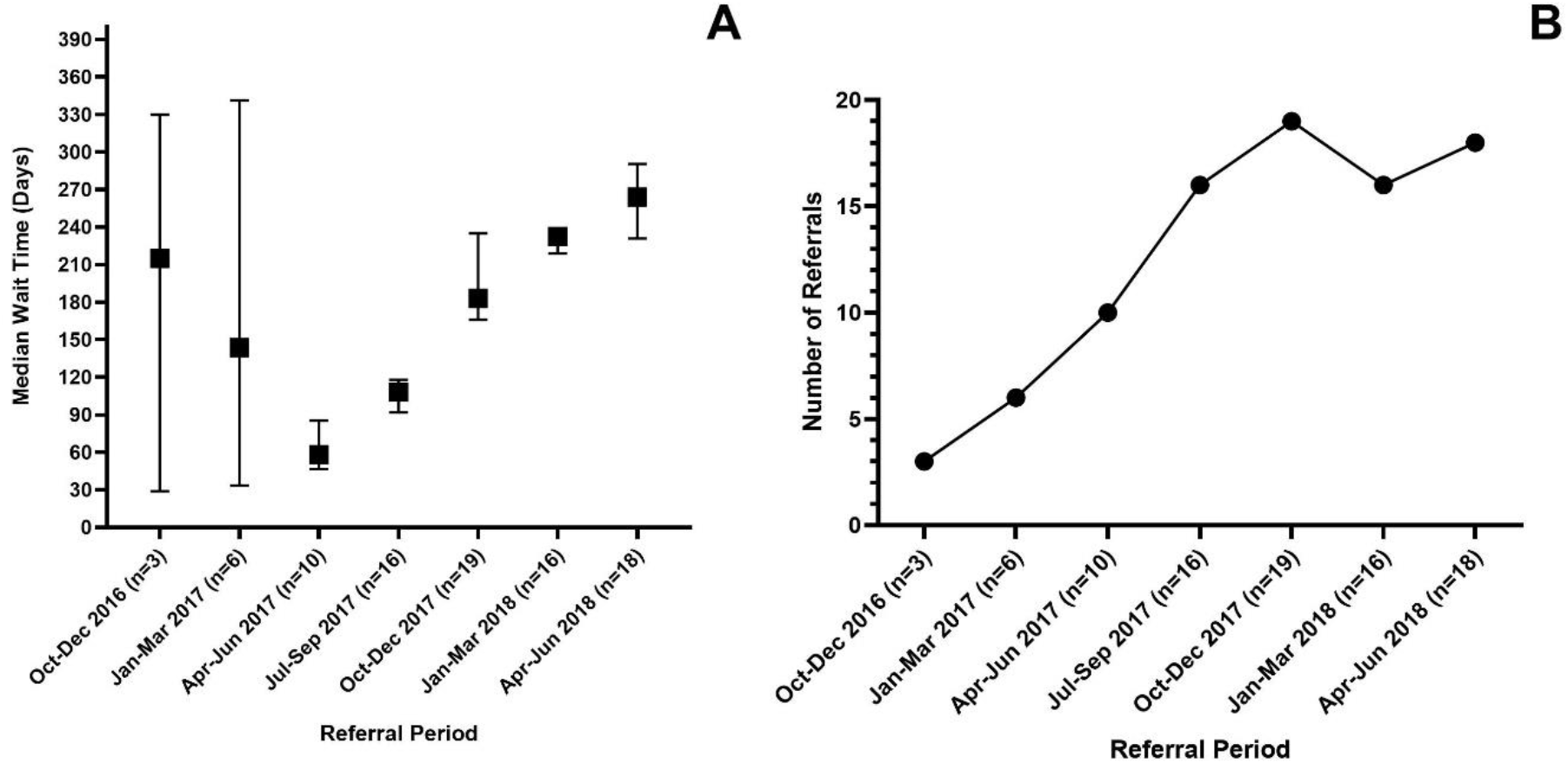
– Referral patterns and wait times. A. Median wait over time (Error bars represent interquartile range) B. Number of referrals over time

The rate of patient retention within the clinic was high. Once seen in KTC, only 7.6% of patients were lost to follow-up. At the time of chart review, the median length of follow-up was 33 weeks [IQR 11-62]. Follow-up was ongoing for 90.2% of patients at the conclusion of this study.

### Quality Indicators

Cervical cancer screening was documented as up to date for 69.2% of eligible people. The remaining 30.8% either did not have screening documented or were late-to-screening (i.e., they did not receive cervical cancer screening in our clinic despite being due to undergo screening).

Of eligible patients, 81.7% had documented fertility preservation offers. Of these, 8/49 patients desired referral to Reproductive Endocrinology and Infertility.

## Discussion

To our knowledge, this is the first Canadian transgender clinic that serves both adults and youth that has described its referral patterns and the services it provides. This clinic is also unique as it is Gynecology-led. The majority of patients were assigned female at birth and more than half identified as transgender male. This gives a ratio of 1:0.44 FtM:MtF. Spanish clinics have described their clinic populations as 1:1.79 FtM:MtF and 1:1. ^11,13^ It is unclear whether the ratio seen in KTC is representative of the population served in Kingston or because the clinic is led by Gynecology.

More than one third of the population referred to KTC were youth patients. This demonstrates that referring providers honour the identities of young patients and recognize the importance of early referral to transgender care. Early referral benefits patients by limiting the irreversible changes that take place in puberty.

Most patients seen at KTC requested hormone therapy, which can be provided by a wide range of providers including family physicians, nurse practitioners, endocrinologists, and gynecologists. There is a compelling argument for transgender health care to be managed by primary care providers as 77.2% of patients sought hormone therapy, which is generally in their scope of practice. Surgery was another common reason for referral. Understanding the funding process and knowing to whom one will refer a patient seeking gender affirmation surgery is an important step in establishing a transgender clinic. Clinicians who consider opening a transgender clinic should note that while referrals give some idea why a patient is presenting – all health providers listed at least one reason for referral that corresponded to the patient’s reasons – only 12% of referrers provide all reasons for which the patient seeks care. This may make referrals challenging to triage.

Our clinic had moderate rates of cervical cancer screening with 69.8% of eligible people documented as up to date on Pap tests. This does not meet Ontario’s cervical cancer screening goal of 85% but exceeds the provincial average screening rate of 65% from 2009-2011. ^20^ The clinic’s late-to-screening rate of 30.2% was on par with the local rate of 22.8-33.7% in 2015. ^21^ This population can be more difficult to screen due to gender dysphoria with pelvic exams and offering self-administered cervical screening or HPV vaginal testing may improve rates. ^27–29^

Our clinic documented fertility preservation offers to 81.7% of eligible patients. The only comparison in the literature is a pediatric and adolescent clinic with offer rates of 98.6%. This will be a target for quality improvement moving forward. It is worth noting that some fertility centres will now offer fertility preservation to transgender men who are already taking testosterone without asking them to pause hormone therapy, though this group was not included in our quality indicator. Of the 49 patients who were offered fertility preservation, 8 (16.3%) desired a referral to REI. This is higher than previously documented rates of 3-12%. ^15,24^ It is unclear if this is due to a bias to offer fertility preservation to those who seem interested or who broach the topic themselves or if the rate would remain as high if all eligible patients were offered fertility preservation. Rate of fertility preservation offer will be a target for quality improvement initiatives.

The limitations of this study include that it was a retrospective chart review without standardized intake forms and was thus subject to reviewer bias and missing data. KTC is run by three physicians and as a teaching centre, histories and documentation were often performed by learners at various stages of training. Where there was missing data the results are subject to non-response bias if cases were removed from analysis. Kingston is a predominantly Caucasian city (10.1% visible minority) and therefore the results of this study may not be applicable to a more diverse population. ^25^

Future research questions include whether the ratio of transgender females to males would differ with more primary care-based centres, whether fertility referral uptake will remain as high if offer rates increase, and how many patients proceed to gamete preservation and live births. We suggest using a standardized intake form immediately at the time of clinic initiation. Data collected in this study would be helpful, as would a patient satisfaction form.

## Conclusion

In the first two years of the Transgender Clinic (KTC) at the Kingston Health Sciences Centre, the referral rates and wait times increased substantially, affirming the need for a clinic of this nature in the Kingston area. Many transgender Kingstonians previously had no gender-affirming care, and some of those who did had to travel hundreds of kilometres for it. Due to the creation of this clinic, they can now have the vast majority of their health needs met locally. This is a standard that could be met in all of Canada by motivated health advocates.

## Data Availability

The data is deidentified patient data and is available through Department of Obstetrics and Gynecology, Queen's University and Kingston Health Sciences Centre. Contact. The data may be used for publication only with the explicit consent of principal investigator Dr. Ashley Waddington.

## Notes

### Competing Interest Statement

The authors have declared no competing interest.

### Funding Statement

No funding was received.

### Author Declarations

Queen's University Health Sciences and Affiliated Teaching Hospitals Research Ethics Board

